# Public Water Quality and Birth Outcomes: Evidence from the World’s Largest Nitrate Removal Facility

**DOI:** 10.1101/2025.05.06.25327112

**Authors:** Jason Semprini

**Affiliations:** Des Moines University, Department of Public Health

## Abstract

In response to rising nitrate levels in Iowa’s public drinking water, the city of Des Moines built the world’s largest nitrate removal facility. The facility operates when nitrate levels exceed the regulatory threshold of 10 mg/L, incurring costs exceeding $10,000 per day. To evaluate the effect of the nitrate removal facility on birth weight and gestational age, we analyzed publicly available birth certificate microdata (1992-2004). In addition to adjusting for maternal and fraternal sociodemographic factors, our linear regression model included county-year fixed-effects to adjust for annual differences across counties and year-month fixed-effects to account for seasonal variation in birth outcomes. Unbiased identification relied on exogenous geotemporal variation in first-trimester exposure to operation of the nitrate removal facility. Operating the nitrate removal facility was associated with a 0.4-percentage-point increase in the probability of normal birth weight and was associated with a 1.6-percentage-point increase in the probability of a normal term birth. The positive associations were largest among mothers with higher risk of adverse outcomes (smoked during pregnancy, cesarean deliveries, prior preterm births, excessive weight gain, unmarried, unknown father). A post-hoc analysis suggests that this intervention may be cost effective, multiplying public investment by 3.5 dollars in health benefits. As policymakers explore how to address rising nitrate pollution, quantifying the value of nitrate reduction strategies could inform future public health interventions. Although the Des Moines nitrate removal facility appears to serve a crucial public health function and advance health equity, challenges remain for deciding how to pay for this potentially cost-effective intervention.

## Introduction

Decades of research have highlighted the adverse relationship between prenatal exposure to water-based nitrate and birth outcomes^1–5^. Regulatory efforts to reduce nitrate pollution have proven ineffective, as evident by the rising incidence of nitrate violations and worsening water quality^6–9^. Instead of leaving nitrate removal responsibilities to individual households, as is common for residents with private well water, some municipal policymakers have invested in nitrate removal systems at public water utilities^10,11^. One example is in Des Moines, Iowa, a state with some of the highest groundwater nitrate in the country^10,12^.

In response to increasing nitrate levels, Des Moines, Iowa’s capitol and largest city, spent $4,100,000 to construct the world’s largest nitrate removal facility^13^. The facility, which costs $10,000 per day of operation, operates when nitrate levels exceed the Environmental Protection Agency (EPA) regulatory threshold of 10 mg/L^13,14^. To date, no research has evaluated how nitrate removal facilities impact health outcomes. As more nitrate removal facilities pop up across the country, the evidence gap muddles the discourse for managing nitrate pollution in America’s increasingly polluted public water.

This evidence is especially needed in predominantly agricultural states where some policymakers pursue developing regional nitrate removal facilities amidst dissenting calls for decentralizing municipal water utilities^15^. Given the high costs of creating and operating nitrate removal facilities, which have been centric to failed litigation and calls for legislative action against groundwater nitrate polluters, research describing the efficacy of the nitrate removal facility can inform the tax-paying public of the nitrate removal facility’s value and allow policymakers at all levels of government to optimize nitrate reduction strategies.

## Methods

### Data and Variables

Seeking to evaluate the impact of Des Moines’ nitrate removal system on birthweight and gestational age outcomes, we accessed 12 years of cross-sectional, publicly available Iowa birth certificate data (1992-2004) from the National Center for Health Statistics (NCHS)^16^. The birth certificate data included all live births, with residential geocodes for births in counties with a population of at least 100,000 people. All births residing in counties with suppressed geocodes were excluded from the study. Note, the NCHS began suppressing all residential geocodes in datafiles 2005-present. The publicly available birth certificate data also included birthweight (grams) and gestational age (weeks), maternal and fraternal sociodemographic variables, and year and month of birth.

For both birthweight and gestational age, birth outcomes were measured as a set of mutually exclusive binary variables. Using the birthweight measure in the birth record, we created mutually exclusive categories for birth weight outcomes: very low birth weight (<1,500 g), low birth weight (1,500-2,499 g), normal birth weight (2,500-4,000 g), high birth weight (>4,000 g). Similarly, we created mutually exclusive categories for gestational age outcomes: very preterm (<32 w), preterm (32-36 w), normal term (37-42 w), and long term (>42 w).

Births with reported birthweight below 500 g and gestational age below 21 weeks were also excluded. All maternal and fraternal sociodemographic variables were dichotomized as a set of categorical control variables measuring marital status, birth order, weight gain, alcohol use, tobacco use, sex of child, delivery, plurality, known father age, mother’s age, mother’s education, prior live birth, prior unalive birth, prior preterm birth. Because date of birth was suppressed, conception date was estimated using birth month/year and gestational age.

The Des Moines nitrate removal facility operation data was obtained via correspondence with staff at Des Moines Water Works (Jeff Mitchell, email correspondence, September 2024). This data identified the number of times the Des Moines nitrate removal facility operated each month since 1991.

### Exposure

Exposure to the nitrate removal facility was based on county of residence and month of conception, as related to operation dates of the Des Moines nitrate removal facility. Births were considered exposed if they both 1) resided in Polk County (FIPS = 19153) and 2) conception occurred between March-July in a year where the Des Moines nitrate removal facility operated >1 day in two consecutive months. Note, the facility never operated between January-February or August-December of any year, and operated intermittently between March-July on each given year.

### Statistical Analysis

Each birth outcome was analyzed via a linear probability regression model. For inference, we estimated standard errors robust to heteroskedasticity and serial autocorrelation, clustered at the county-level^17^. All analyses were conducted in STATA v. 18. In addition to adjusting for maternal and fraternal sociodemographic variables found on a birth certificate, each linear regression model included county-year fixed-effects to adjust for annual differences across counties and year-month fixed-effects to account for seasonal variation in birth outcomes^18,19^.

To assess whether our results were sensitive to analytic decisions and model specifications, we reported the results of multiple sensitivity checks. These sensitivity checks included 1) logistic regression model, 2) linear probability regression model with unclustered standard errors robust to heteroskedasticity and autocorrelation, 3) excluding conception years without consecutive nitrate removal facility operation days >1. For birthweight outcomes, an additional specification included gestational age as an independent variable.

In addition to estimating aggregate effects, we conducted subgroup analyses by maternal sociodemographic and health status characteristics. To assess if estimated effects varied within subgroups, we compared confidence intervals of each estimate, and constructed a t-test for subgroups with two factors or Wald tests for groups with more than two factors.

### Research Design

Our identification strategy relied on geotemporal variation in exposure to operation of the nitrate removal facility. The ideal design for unbiased identification would be a randomized controlled trial, where newly pregnant mothers were randomly assigned to drink water from Des Moines’ nitrate removal facility. Although infeasible retrospectively, such a design would eliminate confounding between potential birth outcomes and unobserved factors correlated with propensity to drink water from the nitrate removal facility. To approximate this ideal with non-experimental observational data, our design leveraged the annual month-to-month variation in nitrate removal facility operation. Equation 1 estimates B1: the association between exposure to operation of the Des Moines nitrate removal facility (D_cmt_) and the change in each modelled outcome (Y_icmtr_).

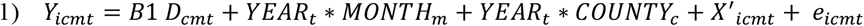

In addition to accounting for observable maternal and fraternal factors (X’_icmt_), Equation 1 also accounted for unobservable annual heterogeneity between counties (YEAR_t_*COUNTY_c_) and temporal trends (YEAR_t_*MONTH_m_) in outcomes. Adjusting for individual birth factors, conditioning on annual differences between each county and temporal trends across the state, and assuming zero residual confounding from within-year seasonal differences between counties, Equation 1 identifies the effect of exposure to the Des Moines Nitrate Removal Facility operation on birth outcomes.

To test the validity of our design and identifying assumption, we implemented a falsification placebo test. Excluding births assigned as exposed in the primary specification, the falsification analysis tested for differences within Polk County births by estimated month of conception: Placebo Exposed = March – July; Unexposed August – February: months when the nitrate removal facility did not actually operate >1 day.

## Results

### Summary Statistics

Between 1992-2004, the Des Moines nitrate removal facility operated for 544 days (Figure 1). Among 48,878 birth records, 171,098 births met the inclusion criteria (Supplemental Exhibit 1). 16,834 births (9.84%) were exposed to operation of the nitrate removal facility (Table 1). Mean birthweight was 3,382.3 grams (g) (SD = 590.9). Mean gestational age was 39.0 weeks (w) (SD = 2.5). 6.4% of the sample was at low birthweight (<2,500 g) and 10.2% of the sample were born preterm (<37 weeks gestation). 81.7% and 85.4% of the sample were normal birthweight and normal term, respectively. Supplemental Exhibits 2-3 report the baseline birth rate outcomes for the full sample and each subgroup population of births in Polk County.

**Table 1:** Birth Record Sample Statistics.

| Variable | Count | Mean | SD |
| --- | --- | --- | --- |
| Birth Weight (g) | 171,098 | 3,382.3* | 590.9 |
| Gestational Age (w) | 171,098 | 39.0* | 2.5 |
| Very Low-Birth Weight (<1500 g) ^ | 1,826 | 0.011 | 0.103 |
| Low-Birth Weight (1500-2500 g) ^ | 9,069 | 0.053 | 0.224 |
| Normal Birth Weight (2500-4000 g) ^ | 139,739 | 0.817 | 0.387 |
| High Birth Weight (>4000 g) ^ | 20,464 | 0.120 | 0.324 |
| Very Preterm Birth (<32 w) ^ | 2,615 | 0.015 | 0.123 |
| Preterm Birth (32-36 w) ^ | 7,558 | 0.044 | 0.205 |
| Normal Term Birth (37-42 w) ^ | 146,139 | 0.854 | 0.353 |
| Long Term Birth (>42 w) ^ | 1,826 | 0.011 | 0.103 |
| Exposed to Operation of Nitrate Removal Facility | 16,834 | 0.098 | 0.298 |
| Teen Pregnancy (age <= 19) | 17,862 | 0.104 | 0.306 |
| High Risk Pregnancy (age >=35) | 17,837 | 0.104 | 0.306 |
| Drank Alcohol During Pregnancy | 3,790 | 0.022 | 0.147 |
| Birth Attended by Provider | 133,711 | 0.781 | 0.413 |
| Smoke During Pregnancy | 32,366 | 0.189 | 0.392 |
| Male Child | 87,540 | 0.512 | 0.500 |
| Vaginal Delivery | 136,878 | 0.800 | 0.400 |
| Plurality Single | 165,974 | 0.970 | 0.170 |
| Mother, no high school degree | 24,547 | 0.143 | 0.351 |
| Mother, high school degree only | 47,641 | 0.278 | 0.448 |
| Mother, college degree | 51,444 | 0.301 | 0.459 |
| non-Hispanic White Mother | 153,190 | 0.895 | 0.306 |
| non-Hispanic Black Mother | 11,857 | 0.069 | 0.254 |
| Known Father Age | 142,441 | 0.833 | 0.373 |
| non-Hispanic White Father | 127,727 | 0.747 | 0.435 |
| Non-Hispanic Black Father | 7,995 | 0.047 | 0.211 |
| Prior Birth Resulting in Death | 3,360 | 0.020 | 0.139 |
| Prior Alive Birth | 100,016 | 0.585 | 0.493 |
| Prior Preterm Birth | 44,549 | 0.260 | 0.439 |
| Mother is Married | 119,176 | 0.697 | 0.460 |
| First Birth | 57,108 | 0.334 | 0.472 |
| Second Birth | 51,024 | 0.298 | 0.457 |
| Third Birth | 31,940 | 0.187 | 0.390 |
| Fourth Birth | 16,292 | 0.095 | 0.294 |
| Fifth Birth | 7,697 | 0.045 | 0.207 |
| Sixth Birth | 3,491 | 0.020 | 0.141 |
| Seventh or more Birth | 3,285 | 0.019 | 0.137 |
| Mother Gained <16 lb | 16,533 | 0.097 | 0.295 |
| Mother Gained 16-20 lb | 16,852 | 0.098 | 0.298 |
| Mother Gained 21-25 b | 23,212 | 0.136 | 0.342 |
| Mother Gained 26-30 lb | 30,270 | 0.177 | 0.382 |
| Mother Gained 31-35 lb | 23,694 | 0.138 | 0.345 |
| Mother Gained 36-40 lb | 21,159 | 0.124 | 0.329 |
| Mother Gained 41-45 lb | 11,328 | 0.066 | 0.249 |
| Mother Gained 46+ lb | 19,963 | 0.117 | 0.321 |

Table 1 reports the summary statistics of the sample of birth records. ^ outcome variable. \* continuous variable. All other variables are binary, with mean values reported on a 0-1 scale. SD = Standard Deviation. g = grams. w = weeks. lb = pounds.

**Table 2:**
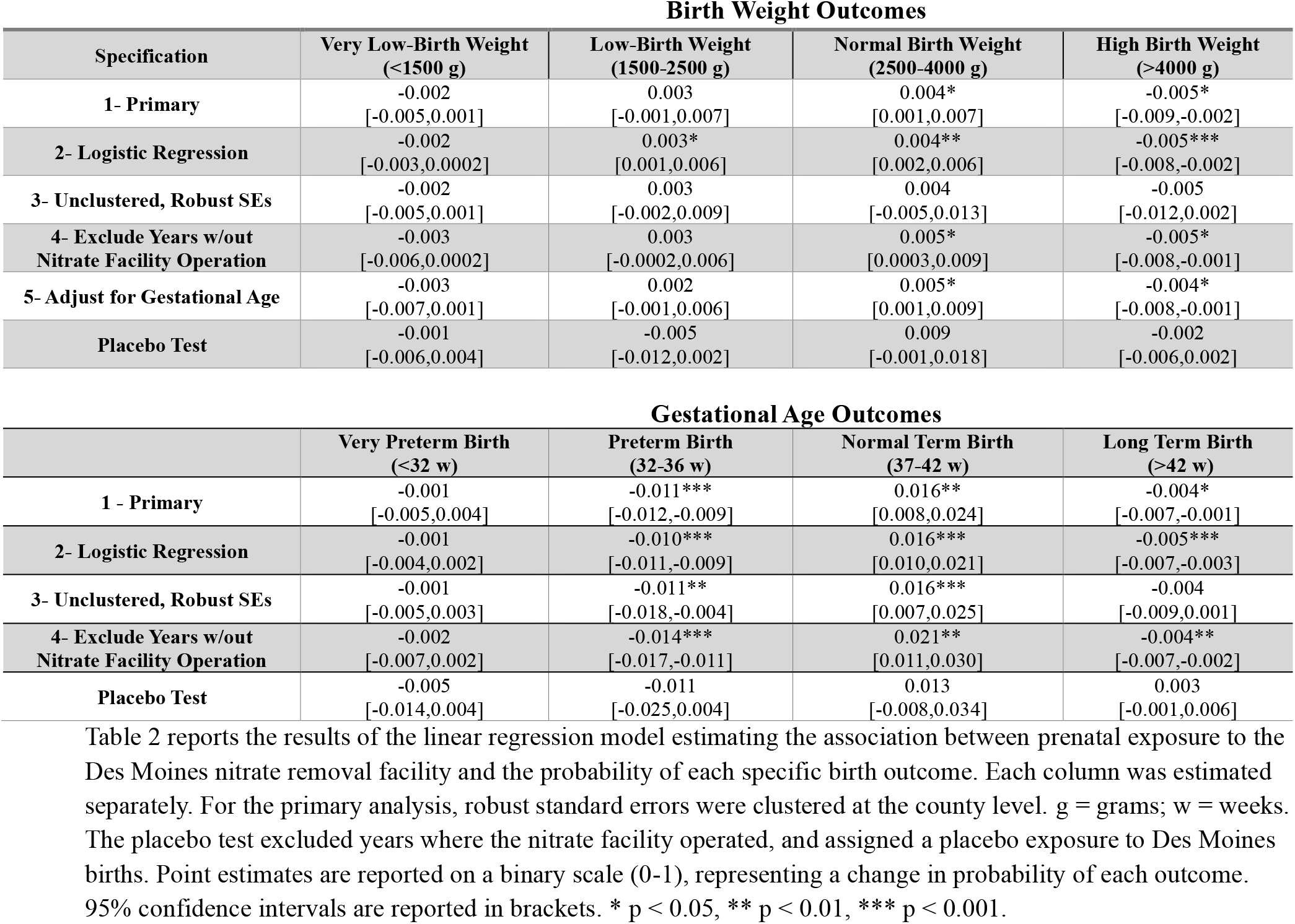
Estimated Associations Between Exposure to Nitrate Removal Facility and Birth Outcomes – Primary Estimates, Sensitivity Checks, and Falsification Tests.

| **Birth Weight Outcomes** |  |  |  |  |
| --- | --- | --- | --- | --- |
| **Specification** | **Very Low-Birth Weight (<1500 g)** | **Low-Birth Weight (1500-2500 g)** | **Normal Birth Weight (2500-4000 g)** | **High Birth Weight (>4000 g)** |
| **1- Primary** | -0.002<br>[-0.005,0.001] | 0.003<br>[-0.001,0.007] | 0.004*<br>[0.001,0.007] | -0.005*<br>[-0.009,-0.002] |
| **2- Logistic Regression** | -0.002<br>[-0.003,0.0002] | 0.003*<br>[0.001,0.006] | 0.004**<br>[0.002,0.006] | -0.005***<br>[-0.008,-0.002] |
| **3- Unclustered, Robust SEs** | -0.002<br>[-0.005,0.001] | 0.003<br>[-0.002,0.009] | 0.004<br>[-0.005,0.013] | -0.005<br>[-0.012,0.002] |
| **4- Exclude Years w/out Nitrate Facility Operation** | -0.003<br>[-0.006,0.0002] | 0.003<br>[-0.0002,0.006] | 0.005*<br>[0.0003,0.009] | -0.005*<br>[-0.008,-0.001] |
| **5- Adjust for Gestational Age** | -0.003<br>[-0.007,0.001] | 0.002<br>[-0.001,0.006] | 0.005*<br>[0.001,0.009] | -0.004*<br>[-0.008,-0.001] |
| **Placebo Test** | -0.001<br>[-0.006,0.004] | -0.005<br>[-0.012,0.002] | 0.009<br>[-0.001,0.018] | -0.002<br>[-0.006,0.002] |

| **Gestational Age Outcomes** |  |  |  |  |
| --- | --- | --- | --- | --- |
|  | **Very Preterm Birth (<32 w)** | **Preterm Birth (32-36 w)** | **Normal Term Birth (37-42 w)** | **Long Term Birth (>42 w)** |
| **1 - Primary** | -0.001<br>[-0.005,0.004] | -0.011***<br>[-0.012,-0.009] | 0.016**<br>[0.008,0.024] | -0.004*<br>[-0.007,-0.001] |
| **2- Logistic Regression** | -0.001<br>[-0.004,0.002] | -0.010***<br>[-0.011,-0.009] | 0.016***<br>[0.010,0.021] | -0.005***<br>[-0.007,-0.003] |
| **3- Unclustered, Robust SEs** | -0.001<br>[-0.005,0.003] | -0.011**<br>[-0.018,-0.004] | 0.016***<br>[0.007,0.025] | -0.004<br>[-0.009,0.001] |
| **4- Exclude Years w/out Nitrate Facility Operation** | -0.002<br>[-0.007,0.002] | -0.014***<br>[-0.017,-0.011] | 0.021**<br>[0.011,0.030] | -0.004**<br>[-0.007,-0.002] |
| **Placebo Test** | -0.005<br>[-0.014,0.004] | -0.011<br>[-0.025,0.004] | 0.013<br>[-0.008,0.034] | 0.003<br>[-0.001,0.006] |

Table 2 reports the results of the linear regression model estimating the association between prenatal exposure to the Des Moines nitrate removal facility and the probability of each specific birth outcome. Each column was estimated separately. For the primary analysis, robust standard errors were clustered at the county level. g = grams; w = weeks. The placebo test excluded years where the nitrate facility operated, and assigned a placebo exposure to Des Moines births. Point estimates are reported on a binary scale (0-1), representing a change in probability of each outcome. 95% confidence intervals are reported in brackets. \* $p < 0.05$ , \*\* $p < 0.01$ , \*\*\* $p < 0.001$ .

**Figure 1:**
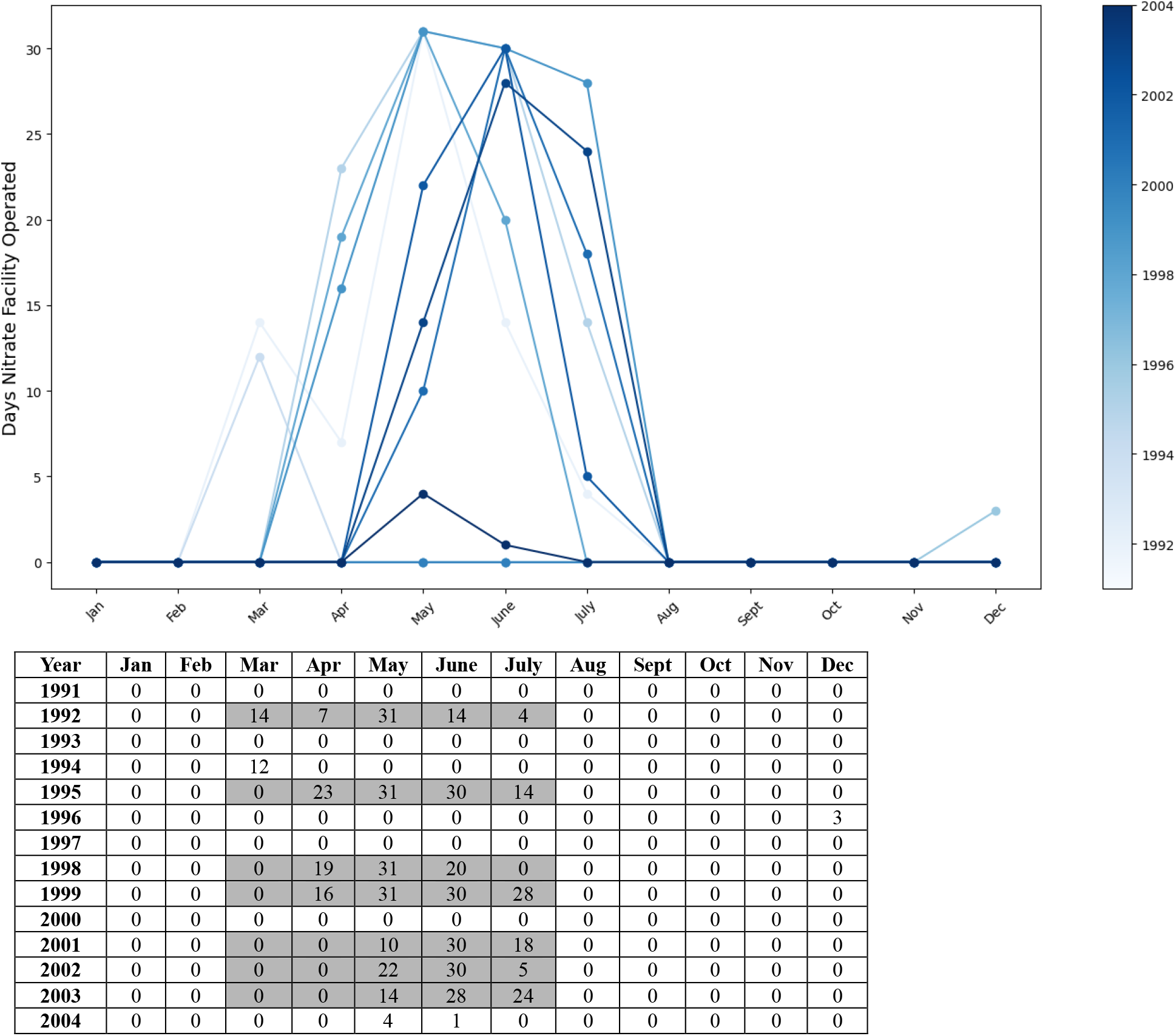
Operation Days for the Des Moines Nitrate Removal Facility (1991-2004) reports the number of days the Des Moines public water nitrate removal facility operated. The facility was constructed and fully operational in 1992. Gray shading indicates births conceived in this month were assigned as exposed to operation of the facility. The placebo test assigned Polk County births where the estimated conception date fell between March-July, for years 1991, 1993, 1994, 1996, 1997, 2000, 2004. *Source: The Des Moines nitrate removal facility operation data was obtained via correspondence with staff at Des Moines Water Works (Jeff Mitchell, email correspondence, September 2024)*.

### Birthweight Outcomes

There was no association between exposure to the facility and the probability of being born at very low birth weight or the probability of being born at low birthweight. There was a statistically significant +0.4-percentage point association between exposure to the nitrate removal facility and normal birth weight (Est. = +0.004, C.I. = .001, 0.007). This represents a 0.5% relative difference from baseline. There was also a statistically significant −0.5-percentage point association between exposure to the facility and high birthweight (Est. = −0.005, C.I. = −0.009, −.002). This estimate represents a 3.8% relative decline from baseline.

Across most alternative specifications, the point estimates and inference were consistent with the primary specification (Table 1). One difference was found in the logistic regression model, where the association between exposure to the nitrate removal facility and probability of low birthweight was statistically significant (Est. = 0.003, C.I. = 0.001, 0.006). For normal birthweight and high birthweight outcomes, the results were consistent across 4/5 alternative specifications, but not when estimating unclustered robust standard errors. For each outcome, results from the falsification test (based on placebo exposure) were not statistically significant.

### Gestational Age Outcomes

There was no association between exposure to the facility and the probability of being very preterm. There was a statistically significant −1.1-percentage point association between exposure to the nitrate removal facility and normal birth weight (Est. = −0.011, C.I. = −0.012, −0.009). This represents a 13.8% relative difference from baseline. There was also a statistically significant +1.6-percentage point association between exposure to the facility and probability of a normal term birth (Est. = 0.016, C.I. = 0.008, 0.024). This estimate represents a 1.9% relative difference from baseline. Finally, there was a statistically significant −0.4-percentage point association between exposure to the nitrate removal facility and long term birth (Est. = −0.004, C.I. = −0.007, −0.001). This represents a 5.4% relative difference from baseline.

For very preterm, preterm, and normal term birth outcomes, the results from each of the five alternative specifications were consistent with the primary results (Table 1). For long term birth outcomes, only the specification estimating unclustered robust standard errors diverged from the inference of the primary specification. None of the falsification test estimates were statistically significant.

### Subgroup Analyses

#### Birthweight Outcomes

Figure 2 reports the estimated association between early prenatal exposure to the nitrate removal facility and the probability of normal birthweight, across subgroup populations. See supplemental exhibit 4 for the reported estimates for each birthweight category. In summary, the association between exposure to the facility and normal birthweight did not vary by age group, birth attendant status, child sex, delivery method, maternal race, known father’s age, or maternal marital status. The associations between exposure to the facility and normal birth weight were largest for mothers who smoked during pregnancy (Est. = 0.009, C.I. = 0.005, 0.013) and (Est. = 0.023, C.I. = 0.013, 0.033).

**Figure 2:**
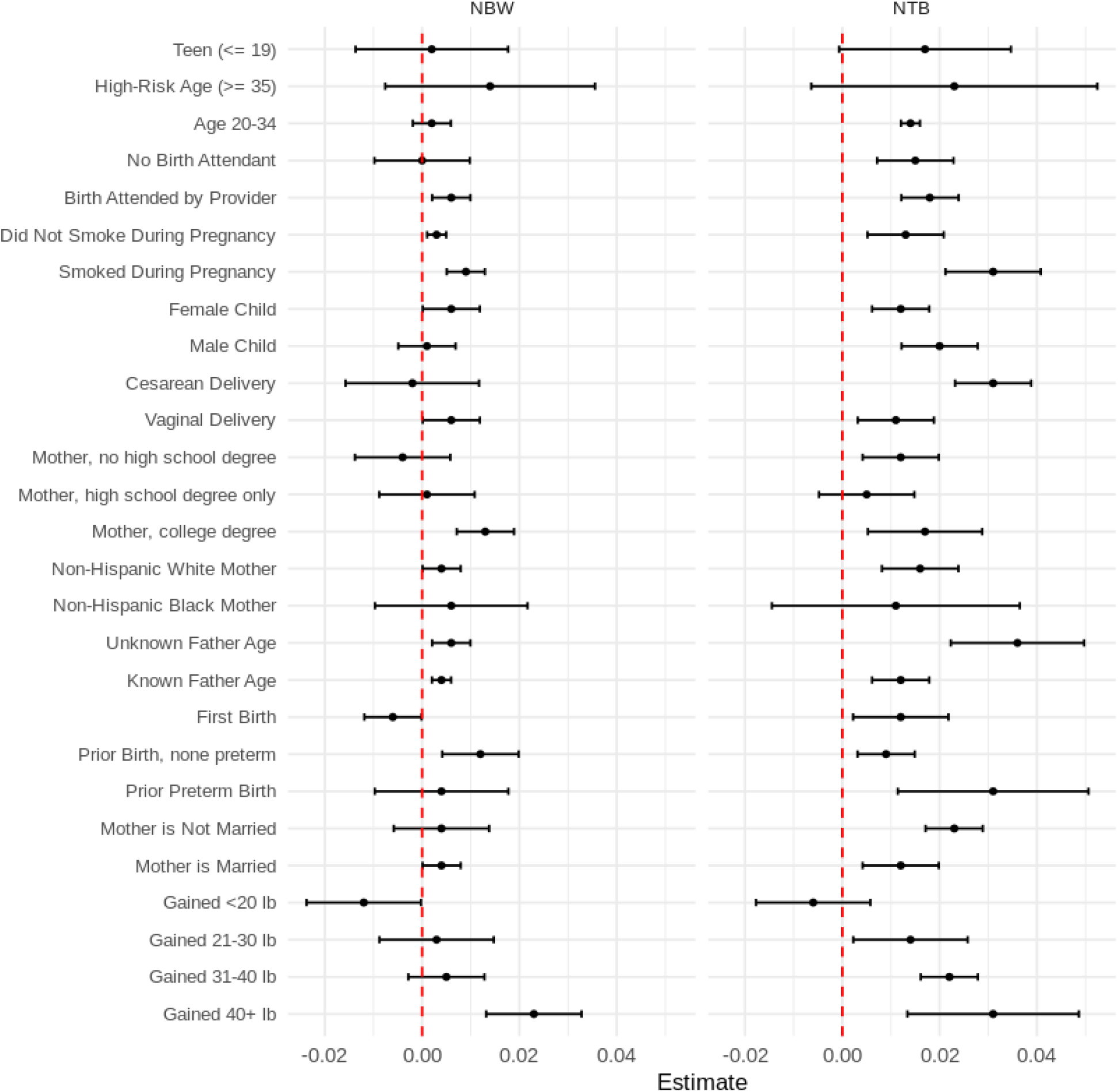
Subgroup Results for Normal Birthweight and Normal Term Birth Outcomes. visualizes the subgroup estimates for the primary linear regression model specification, estimating the association between prenatal exposure to the Des Moines nitrate removal facility and the probability of 1) normal birth weight (NBW) and 2) normal term birth (NTB). Normal birth weight = 2500-4000 grams. Normal term birth = 37-42 weeks gestational age. Robust standard errors were clustered at county level. Error bars represent 95% confidence interval of the point estimate. Point estimates are reported on a binary scale (0-1), representing a change in probability of each outcome.

There were statistically significant differences across maternal smoking status, where the association was smaller in mothers who didn’t smoke during pregnancy (Est. = 0.003, C.I. = 0.001, 0.004) than the association in mothers who did smoke during pregnancy (Est. = 0.009, C.I. = 0.005, 0.013). In mothers who smoked during pregnancy, exposure to the nitrate removal facility was also associated with a 0.6-percentage point decline in the probability of being born at very low birthweight (Est. = −0.006, C.I. = −0.010, −0.002; Supplemental Exhibit 4).

The association between exposure to the facility and normal birthweight also varied by maternal education status. Estimated associations were near zero and statistically insignificant for mothers without a high school degree and mothers with a high school degree only. In mothers with a college degree, exposure to the facility was associated with a +1.3-percentage-point change in the probability of normal birthweight (Est. = 0.013, C.I. = 0.007, 0.019).

Finally, the estimated associations varied by birth history and weight gain during the current pregnancy. While the association was negative for first births (Est. = −0.006, C.I. = −0.012, −0.0001), the association was positive for mothers with prior births which were not preterm (Est. = 0.012, C.I. = 0.004, 0.020; Supplemental Exhibit 4). The association was null for mothers with a prior preterm birth. For mothers which gained less than 16 lb during pregnancy, exposure was negatively (Est. = −0.012, C.I. = −0.024, −0.0002) and for mothers which gained more than 40 lb during pregnancy exposure was positively (Est. = 0.023, C.I. = 0.013, 0.033) associated with normal birth weight.

#### Gestational Age Outcomes

Figure 2 reports the estimated association between early prenatal exposure to the nitrate removal facility and the probability of normal term births, across subgroup populations. See supplemental exhibit 4 for the reported estimates for each gestational age category. In summary, the association between exposure to the facility and normal term birth did not vary by age group, birth attendant status, child sex, maternal education, and maternal race. Like the results for birthweight, there were statistically significant differences across maternal smoking status, birth history, and weight gain during pregnancy. Similarly, the association between exposure to the facility and normal term birth was largest for mothers who smoked during pregnancy (Est. = 0.031, C.I. = 0.021, 0.041) and mothers who gained > 40 lb during pregnancy. (Est. = 0.031, C.I. = 0.013, 0.049). Additionally, associations with normal term births were largest among caesarean-section births (Est. = 0.031, C.I. = 0.023, 0.039), mothers with a prior preterm birth (Est. = 0.031, C.I. = 0.011, 0.051), unmarried mothers (Est. = 0.023, C.I. = 0.017, 0.029), and births with no information on the father (Est. = 0.036, C.I. = 0.022, 0.050),

The association was smaller in mothers who didn’t smoke during pregnancy (Est. = 0.0.13, C.I. = 0.005, 0.021) than the association in mothers who did smoke during pregnancy (Est. = 0.031, C.I. = 0.021, 0.041). In mothers who smoked during pregnancy, exposure to the nitrate removal facility was also associated with a 2.2-percentage point decline in the probability of being born preterm (Est. = −0.022, C.I. = −0.028, −0.016; Supplemental Exhibit 6); a 28.2% relative reduction from baseline. Again, like the estimates for birthweight, the association for normal term birth was lowest for pregnancies gaining < 16 lb (Est. = −0.006, C.I. = −0.018, 0.006) and highest for pregnancies gaining > 40 lb (Est. = 0.031, C.I. = 0.013, 0.049).

The association between exposure to the facility and normal birthweight also varied by maternal marital status and whether the father’s age was known. The association was smaller in married mothers (Est. = 0.012, C.I. = 0.004, 0.019) than the respective association in unmarried mothers (Est. = 0.023, C.I. = 0.017, 0.029; t-test statistic p=0.028). The association was smaller in births with information about the father’s age (Est. = 0.012, C.I. = 0.006, 0.018) than the association in births without information about the father’s age (Est. = 0.036, C.I. = 0.022, 0.050). In both births from unmarried mothers and births with unknown father age information, exposure to the facility was associated with declines in the probability of preterm births; representing, respectively, a 16% and 30% relative change from baseline preterm rates.

Unlike the results for birthweight, the association between exposure to the facility and normal term birth varied by delivery method and prior preterm birth history. The estimated association was smaller in vaginal delivery births (Est. = 0.011, C.I. = 0.003, 0.019) than cesarean delivery births (Est. = 0.031, C.I. = 0.023, 0.039). For cesarean births, the 2.2-percentage point association between exposure to the nitrate removal facility and probability of preterm birth represents a 20.0% relative reduction from baseline (Supplemental Exhibit 4). Finally, while the associations for first births and births without a history of preterm delivery were positive and statistically significant, the largest association was found in births where the mother had delivered a preterm birth in the past (Est. = 0.031, C.I. = 0.011, 0.051). For mothers with a prior preterm birth, the 2.3-percentage point association between exposure to the nitrate removal facility and probability of preterm birth represents a 26.7% relative reduction from baseline (Supplemental Exhibit 4).

## Discussion

Focusing on a metropolitan community at the center of an agricultural state with high, rising levels of groundwater nitrate, our analysis suggests that the world’s largest nitrate removal facility serves a critical public health function while advancing health equity goals^20^. Based on size and consistency across model specifications, the strongest evidence for the nitrate removal facility’s protective impact was related to reducing the probability of preterm births. Despite differences in geographies and settings, our results are quite consistent with the most rigorous existing evidence quantifying the adverse consequences of exposure to prenatal nitrate^21–24^. Although, to be clear, birth outcomes would have been best under a scenario with zero exposure to water-based nitrate^21,23^. Still, if the Des Moines nitrate removal facility never operated, hundreds of more babies may have been born at unhealthy weights or gestational ages.

While this study did not investigate any outcomes beyond birth status, the implications of a public health intervention which effectively improves the likelihood of normal term and normal weight births extends far beyond the delivery room. For decades, research has shown how term and birth weight are not just correlated with, but predictive of neonatal complications and developmental delays, as well as disability and mortality during infancy^25– 29^. The impact of poor birth outcomes can extend across the life course, as evident by research showing how ten-year survival in very preterm births was 66%, compared to 99% in normal term births^30^. Even in adulthood, a recent review from decades of preterm birth cohort studies suggests that survivors of poor birth outcomes have increased probability of cardiovascular, metabolic, respiratory, psychiatric, and neurodevelopmental diseases^31^. With risks elevating at earlier gestational ages, adults born preterm sustained 30-50% higher risk of early adult or midlife mortality than comparable peers born at full term^31^. While much is known about the long-term consequences of poor birth outcomes, less is known about the long-term benefits and costs from interventions which promote healthy birth outcomes. Building off this current study, future research linking birth certificates to medical or vital records can examine health outcomes twenty to thirty years after exposure to nitrate removal facilities.

Public health interventions cannot be evaluated by benefits alone. Costs must also be considered. During the study period, Des Moines’ nitrate removal facility operated for 544 days at a cost of $10,000 per day^13^. Including the $4.1 million construction costs, by the end of 2004, operating the nitrate removal facility cost $9,540,000…all of which was paid with tax dollars from Des Moines residents. A crude, post-hoc cost analysis suggests that the $9,540,000 cost of operating the nitrate removal facility was associated with 676 additional births with healthy weight and gestational age. By operating the nitrate removal facility, the Des Moines’ residents paid $14,112 for each additional healthy birth.

Interpreted causally, the Des Moines’ nitrate removal facility appears to be an extremely cost-effective intervention for promoting healthy births. Consider first, that, on average compared to full term births, each preterm birth incurs an additional $36,000 in first year medical expenses^32^. This would suggest a 2-1 benefit to cost ratio for the facility. Then, after considering additional medical, educational, or social support expenses beyond the first year, the total cost exceeds $50,000 for each preterm birth^33^. By potentially averting these adverse birth outcomes, the nitrate removal facility generates $33,800,000 in public benefits. In summary, every public dollar spent operating the nitrate removal facility yields a 3.5 benefit/cost multiplier. The question then becomes, *who* should pay for this cost-effective intervention?

Cost-benefit analyses can help us decide, as a society, if we should implement a public health intervention. While validation and longer term research will be needed to build upon this current study, this evidence clearly shows that the public should implement the nitrate removal facility. Yet, a cost-benefit analysis provides no informative guidance on who should incur the cost of public health interventions. A utilitarian public health equity perspective may argue that the Des Moines residents should continue spending tax revenue, as evident by both the generalizable benefits for all and elevated benefits for some vulnerable populations^34,35^. However, the benefits from the intervention would also extend beyond the local population due to reduced health system costs. This is especially relevant for birth outcomes, given that Medicaid pays for nearly half of all preterm births^36^. As Medicaid service delivery shifts towards value-based, managed care amidst a growing interest in addressing social determinants of health among enrollees, an argument could be made that state Medicaid programs should shoulder some of the costs of operating the nitrate removal facility^34,37–39^.

These arguments may prove unsatisfactory, especially for free market enthusiasts who would prefer to address the externality of nitrate pollution more directly^40–43^. An externality is a societal cost that is not included in the price of a good or service. In this context, agricultural producers do not include the cost of nitrate pollution in the price of agricultural products. Here, nitrate pollution carries an externality in the form of adverse birth outcomes. Addressing the externality of nitrate pollution, thus, would require considering the role of polluters. A classic free market perspective, then, would argue for taxing the nitrate polluters an amount “equivalent to the cost of harm to others”^44,45^. An alternative libertarian perspective would argue that all a society needs to address externalities are effective governance structures protecting property rights; which would in turn allow people harmed or infringed by nitrate pollution to seek compensation directly from the polluter through judicial processes^46^.

Deciding who should pay the cost of Des Moines’ nitrate removal facility lies far outside the scope of this study. We simply offered a few perspectives to stimulate debate. Recent trends suggest that policymakers across the globe will be forced to respond, in some way or another, to the rising levels of nitrate in our water^47^. Whether society responds to rising nitrate levels by deciding to construct and operate nitrate removal facilities may be of less consequence for promoting healthy births than deciding who should pay for protecting mothers from this harmful chemical.

## Limitations

This study has several limitations. First, the data only included births from metropolitan counties in a small agricultural state in the Midwest during the 1990’s-2000’s. Whether these findings generalize beyond these specific places and times requires future research. Second, because the actual dates of birth were suppressed, the design relied on month and year of birth to estimate conception dates, potentially misclassifying exposure. The design aimed to minimize the threat of this limitation by relying on broad range of exposure based on operation of the nitrate removal facility. Both of these limitations can and should be addressed with research on other states, at different time periods, with restricted data. Third, despite rigorous adjustment with county-year and year-month fixed effects, residual confounding from unobserved factors correlated with both nitrate exposure and birth outcomes which vary across counties differently each year (i.e., a flood disaster/emergency) could potentially bias these estimates. While the placebo tests help validate the design, no empirical tests can fully confirm that the design is unbiased. Therefore, all causal interpretations should consider this context.

## Conclusion

In response to rising nitrate levels in Iowa’s public drinking water, in 1992 the city of Des Moines built the world’s largest nitrate removal facility. Including construction and daily operation expenses between 1992-2004, the facility cost Des Moines taxpayers over $14 million. By linking daily nitrate removal facility operation data with public birth records, this study evaluated the association between exposure to operation of the Des Moines nitrate removal facility and birth outcomes (weight, gestational age). The research design accounted for unobserved confounding that varied annually between counties and unobserved seasonal variation consistent across the state. The identification strategy relied on exogenous geotemporal variation in first-trimester exposure to operation of the nitrate removal facility. The results suggest that operating the nitrate removal facility was associated with a 0.4-percentage-point increase in the probability of normal birth weight and a 1.6%-point increase in the probability of a normal term birth. These positive associations were largest among mothers at higher risks for adverse outcomes. As lawmakers debate how to improve public water quality amidst rising agricultural pollution, identifying and quantifying the value of nitrate reduction strategies could inform nitrate reduction strategies. Validation studies and research evaluating longer term outcomes, outside the state of Iowa could further inform whether nitrate removal facilities should be scaled regional. Although this early evidence suggests that the Des Moines nitrate removal facility serves a critical public health role and may advance health equity, questions remain about who should pay for this potentially cost-effective intervention.

## Supporting information

Supplemental Exhibits

## Data Availability

Data sharing is prohibited by third-party restrictions.

