## Supplemental Exhibits for "Public Water Quality and Birth Outcomes: Evidence from the World’s Largest Nitrate Removal Facility"

**Supplemental Exhibit**

**Supplemental Exhibit 1: Inclusion Criteria**

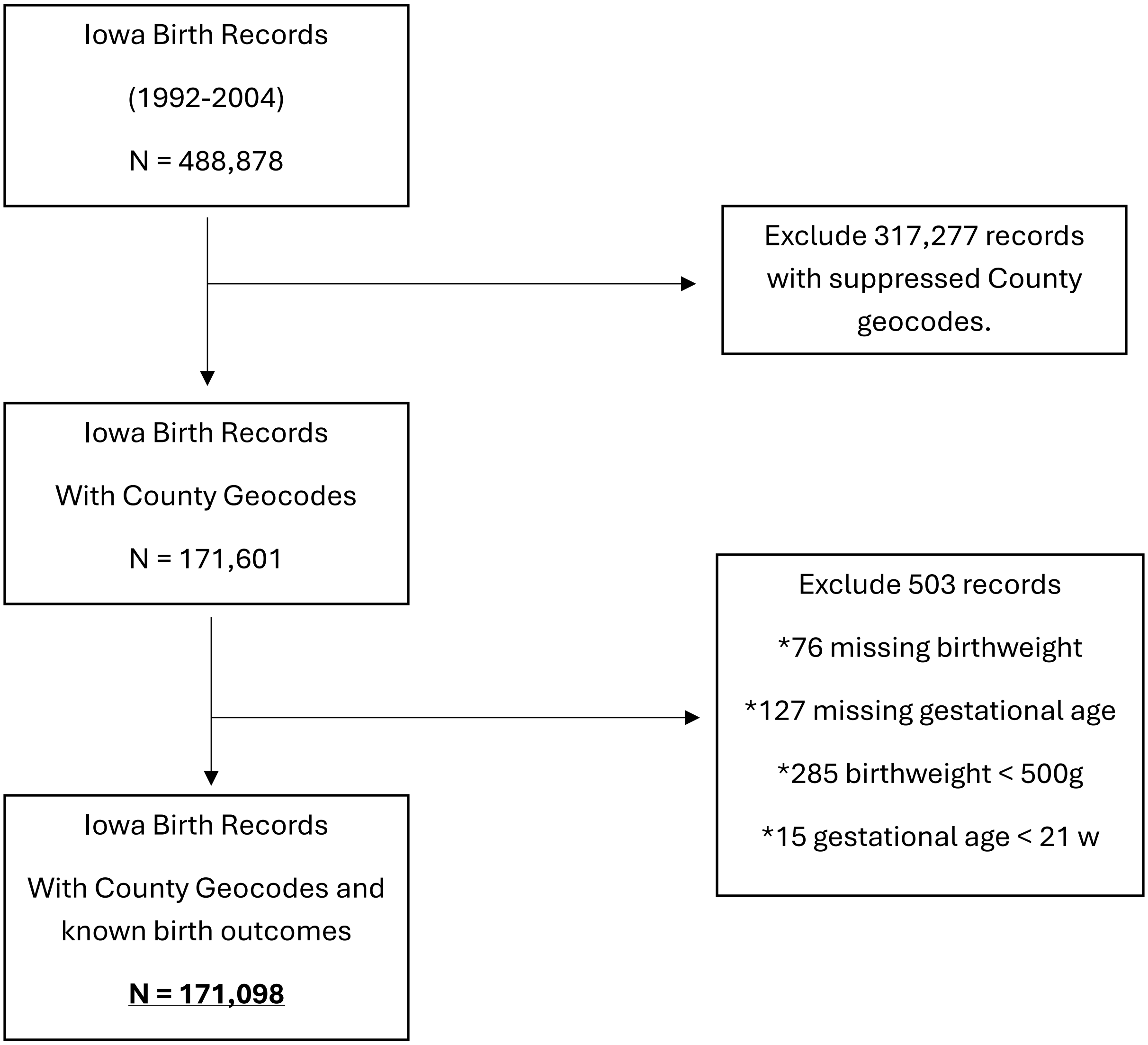

### Supplemental Exhibit 2: Baseline Birthweight Rates for Polk County

### (1991-1992 Conception Year)

| Group | Very Low-Birth Weight (<1500 g) | Low-Birth Weight (1500-2500 g) | Normal Birth Weight (2500-4000 g) | High Birth Weight (>4000 g) |
|---|---|---|---|---|
| Total Sample | 0.009 | 0.051 | 0.806 | 0.133 |
| Teen Pregnancy (age <=19) | 0.019 | 0.060 | 0.845 | 0.077 |
| High-risk Pregnancy (age >=35) | 0.005 | 0.054 | 0.763 | 0.178 |
| Mother Age 20-34 | 0.009 | 0.050 | 0.805 | 0.136 |
| No Birth Attendant | 0.012 | 0.061 | 0.811 | 0.116 |
| Birth Attended by Provider | 0.009 | 0.047 | 0.804 | 0.140 |
| Did Not Smoke During Pregnancy | 0.008 | 0.044 | 0.794 | 0.154 |
| Smoked During Pregnancy | 0.013 | 0.078 | 0.853 | 0.056 |
| Female Child | 0.009 | 0.057 | 0.834 | 0.100 |
| Male Child | 0.010 | 0.046 | 0.779 | 0.165 |
| Cesarean Delivery | 0.026 | 0.082 | 0.709 | 0.183 |
| Vaginal Delivery | 0.005 | 0.044 | 0.831 | 0.121 |
| Mother, no high school degree | 0.021 | 0.071 | 0.832 | 0.075 |
| Mother, high school degree only | 0.011 | 0.064 | 0.802 | 0.123 |
| Mother, college degree | 0.002 | 0.034 | 0.787 | 0.177 |
| Non-Hispanic White Mother | 0.009 | 0.049 | 0.801 | 0.141 |
| Non-Hispanic Black Mother | 0.023 | 0.071 | 0.842 | 0.065 |
| Unknown Father Age | 0.021 | 0.073 | 0.811 | 0.095 |
| Known Father Age | 0.005 | 0.044 | 0.805 | 0.146 |
| First Birth | 0.009 | 0.058 | 0.820 | 0.113 |
| Prior Birth, none preterm | 0.008 | 0.045 | 0.798 | 0.149 |
| Prior Preterm Birth | 0.011 | 0.052 | 0.801 | 0.136 |
| Mother is Not Married | 0.021 | 0.071 | 0.813 | 0.095 |
| Mother is Married | 0.005 | 0.044 | 0.803 | 0.148 |
| Gained <20 lb | 0.015 | 0.094 | 0.810 | 0.080 |
| Gained 21-30 lb | 0.007 | 0.046 | 0.841 | 0.107 |
| Gained 31-40 lb | 0.005 | 0.038 | 0.814 | 0.144 |
| Gained 40+ lb | 0.007 | 0.028 | 0.728 | 0.237 |

Birthweight outcomes are classified as a set of mutually exclusive groups. Rates are reported on a binary scale (0-1), reflecting the proportion of the analytic sample.

### Supplemental Exhibit 3: Baseline Gestational Age Rates for Polk County

### (1991-1992 Conception Year)

| Group | Very Preterm Birth (<32 w) | Preterm Birth (32-36 w) | Normal Term Birth (37-42 w) | Long Term Birth (>42 w) |
|---|---|---|---|---|
| Total Sample | 0.012 | 0.080 | 0.834 | 0.074 |
| Teen Pregnancy (age <=19) | 0.022 | 0.109 | 0.779 | 0.090 |
| High-risk Pregnancy (age >=35) | 0.007 | 0.091 | 0.849 | 0.052 |
| Mother Age 20-34 | 0.011 | 0.075 | 0.839 | 0.075 |
| No Birth Attendant | 0.014 | 0.095 | 0.804 | 0.087 |
| Birth Attended by Provider | 0.012 | 0.073 | 0.846 | 0.069 |
| Did Not Smoke During Pregnancy | 0.011 | 0.078 | 0.843 | 0.068 |
| Smoked During Pregnancy | 0.018 | 0.086 | 0.799 | 0.097 |
| Female Child | 0.013 | 0.076 | 0.833 | 0.078 |
| Male Child | 0.012 | 0.083 | 0.835 | 0.071 |
| Cesarean Delivery | 0.023 | 0.110 | 0.797 | 0.070 |
| Vaginal Delivery | 0.009 | 0.072 | 0.843 | 0.075 |
| Mother, no high school degree | 0.026 | 0.107 | 0.779 | 0.087 |
| Mother, high school degree only | 0.016 | 0.083 | 0.810 | 0.091 |
| Mother, college degree | 0.002 | 0.070 | 0.874 | 0.054 |
| Non-Hispanic White Mother | 0.011 | 0.077 | 0.837 | 0.076 |
| Non-Hispanic Black Mother | 0.026 | 0.126 | 0.790 | 0.058 |
| Unknown Father Age | 0.028 | 0.099 | 0.783 | 0.090 |
| Known Father Age | 0.007 | 0.073 | 0.851 | 0.069 |
| First Birth | 0.012 | 0.092 | 0.823 | 0.073 |
| Prior Birth, none preterm | 0.012 | 0.064 | 0.846 | 0.078 |
| Prior Preterm Birth | 0.013 | 0.086 | 0.831 | 0.071 |
| Mother is Not Married | 0.029 | 0.099 | 0.783 | 0.089 |
| Mother is Married | 0.006 | 0.072 | 0.853 | 0.069 |
| Gained <20 lb | 0.023 | 0.106 | 0.796 | 0.075 |
| Gained 21-30 lb | 0.007 | 0.076 | 0.848 | 0.069 |
| Gained 31-40 lb | 0.009 | 0.071 | 0.847 | 0.073 |
| Gained 40+ lb | 0.008 | 0.061 | 0.852 | 0.080 |

Gestational age outcomes are classified as a set of mutually exclusive groups. Rates are reported on a binary scale (0-1), reflecting the proportion of the analytic sample.

### Supplemental Exhibit 4: Subgroup Estimates, All Outcomes

| Subgroup | Outcome | Est | se | Lower | Upper | Category |
|---|---|---|---|---|---|---|
| No Birth Attendant | VPTB | 0.001 | 0.002 | -0.00292 | 0.00492 | Gestational Age |
| Birth Attended by Provider | VPTB | -0.001 | 0.002 | -0.00492 | 0.00292 | Gestational Age |
| Did Not Smoke During Pregnancy | VPTB | 0 | 0.002 | -0.00392 | 0.00392 | Gestational Age |
| Smoked During Pregnancy | VPTB | -0.004 | 0.003 | -0.00988 | 0.00188 | Gestational Age |
| Female Child | VPTB | -0.002 | 0.002 | -0.00592 | 0.00192 | Gestational Age |
| Male Child | VPTB | 0.001 | 0.001 | -0.00096 | 0.00296 | Gestational Age |
| Cesarean Delivery | VPTB | -0.005 | 0.005 | -0.0148 | 0.0048 | Gestational Age |
| Vaginal Delivery | VPTB | 0 | 0.001 | -0.00196 | 0.00196 | Gestational Age |
| Mother, no high school degree | VPTB | 0 | 0.002 | -0.00392 | 0.00392 | Gestational Age |
| Mother, high school degree only | VPTB | 0.002 | 0.001 | 4E-05 | 0.00396 | Gestational Age |
| Mother, college degree | VPTB | -0.006 | 0.005 | -0.0158 | 0.0038 | Gestational Age |
| Non-Hispanic White Mother | VPTB | -0.001 | 0.002 | -0.00492 | 0.00292 | Gestational Age |
| Non-Hispanic Black Mother | VPTB | 0.001 | 0.005 | -0.0088 | 0.0108 | Gestational Age |
| Unknown Father Age | VPTB | 0.001 | 0.003 | -0.00488 | 0.00688 | Gestational Age |
| Known Father Age | VPTB | -0.001 | 0.002 | -0.00492 | 0.00292 | Gestational Age |
| First Birth | VPTB | 0.005 | 0.001 | 0.00304 | 0.00696 | Gestational Age |
| Prior Birth, none preterm | VPTB | -0.003 | 0.003 | -0.00888 | 0.00288 | Gestational Age |
| Prior Preterm Birth | VPTB | -0.005 | 0.003 | -0.01088 | 0.00088 | Gestational Age |
| Mother is Not Married | VPTB | 0 | 0.003 | -0.00588 | 0.00588 | Gestational Age |
| Mother is Married | VPTB | -0.001 | 0.001 | -0.00296 | 0.00096 | Gestational Age |
| Gained <20 lb | VPTB | 0.003 | 0.002 | -0.00092 | 0.00692 | Gestational Age |
| Gained 21-30 lb | VPTB | -0.003 | 0.002 | -0.00692 | 0.00092 | Gestational Age |
| Gained 31-40 lb | VPTB | -0.005 | 0.003 | -0.01088 | 0.00088 | Gestational Age |
| Gained 40+ lb | VPTB | 0.001 | 0.001 | -0.00096 | 0.00296 | Gestational Age |
| Teen (<= 19) | VPTB | 0 | 0.004 | -0.00784 | 0.00784 | Gestational Age |
| High-Risk Age (>= 35) | VPTB | -0.001 | 0.007 | -0.01472 | 0.01272 | Gestational Age |
| Age 20-34 | VPTB | -0.001 | 0.001 | -0.00296 | 0.00096 | Gestational Age |
| No Birth Attendant | VLBW | 0 | 0.001 | -0.00196 | 0.00196 | Birthweight |
| Birth Attended by Provider | VLBW | -0.002 | 0.002 | -0.00592 | 0.00192 | Birthweight |
| Did Not Smoke During Pregnancy | VLBW | -0.001 | 0.001 | -0.00296 | 0.00096 | Birthweight |
| Smoked During Pregnancy | VLBW | -0.006 | 0.002 | -0.00992 | -0.00208 | Birthweight |
| Female Child | VLBW | -0.004 | 0.002 | -0.00792 | -8E-05 | Birthweight |
| Male Child | VLBW | 0.001 | 0.002 | -0.00292 | 0.00492 | Birthweight |
| Cesarean Delivery | VLBW | -0.001 | 0.004 | -0.00884 | 0.00684 | Birthweight |
| Vaginal Delivery | VLBW | -0.002 | 0.001 | -0.00396 | -4E-05 | Birthweight |
| Mother, no high school degree | VLBW | -0.003 | 0.002 | -0.00692 | 0.00092 | Birthweight |
| Mother, high school degree only | VLBW | 0 | 0.002 | -0.00392 | 0.00392 | Birthweight |
| Mother, college degree | VLBW | -0.004 | 0.004 | -0.01184 | 0.00384 | Birthweight |
| Non-Hispanic White Mother | VLBW | -0.002 | 0.001 | -0.00396 | -4E-05 | Birthweight |
| Non-Hispanic Black Mother | VLBW | -0.004 | 0.004 | -0.01184 | 0.00384 | Birthweight |
| Unknown Father Age | VLBW | -0.006 | 0.002 | -0.00992 | -0.00208 | Birthweight |

| | | | | | | |
|---|---|---|---|---|---|---|
| Known Father Age | VLBW | -0.001 | 0.001 | -0.00296 | 0.00096 | Birthweight |
| First Birth | VLBW | 0.002 | 0.001 | 4E-05 | 0.00396 | Birthweight |
| Prior Birth, none preterm | VLBW | -0.003 | 0.002 | -0.00692 | 0.00092 | Birthweight |
| Prior Preterm Birth | VLBW | -0.004 | 0.004 | -0.01184 | 0.00384 | Birthweight |
| Mother is Not Married | VLBW | -0.006 | 0.002 | -0.00992 | -0.00208 | Birthweight |
| Mother is Married | VLBW | 0 | 0.001 | -0.00196 | 0.00196 | Birthweight |
| Gained <20 lb | VLBW | 0.001 | 0.002 | -0.00292 | 0.00492 | Birthweight |
| Gained 21-30 lb | VLBW | -0.004 | 0.003 | -0.00988 | 0.00188 | Birthweight |
| Gained 31-40 lb | VLBW | -0.004 | 0.002 | -0.00792 | -8E-05 | Birthweight |
| Gained 40+ lb | VLBW | 0.002 | 0.001 | 4E-05 | 0.00396 | Birthweight |
| Teen (<= 19) | VLBW | -0.007 | 0.004 | -0.01484 | 0.00084 | Birthweight |
| High-Risk Age (>= 35) | VLBW | 0.004 | 0.004 | -0.00384 | 0.01184 | Birthweight |
| Age 20-34 | VLBW | -0.002 | 0.001 | -0.00396 | -4E-05 | Birthweight |
| No Birth Attendant | PTB | -0.011 | 0.004 | -0.01884 | -0.00316 | Gestational Age |
| Birth Attended by Provider | PTB | -0.013 | 0.001 | -0.01496 | -0.01104 | Gestational Age |
| Did Not Smoke During Pregnancy | PTB | -0.008 | 0.001 | -0.00996 | -0.00604 | Gestational Age |
| Smoked During Pregnancy | PTB | -0.022 | 0.003 | -0.02788 | -0.01612 | Gestational Age |
| Female Child | PTB | -0.006 | 0.002 | -0.00992 | -0.00208 | Gestational Age |
| Male Child | PTB | -0.015 | 0.003 | -0.02088 | -0.00912 | Gestational Age |
| Cesarean Delivery | PTB | -0.023 | 0.003 | -0.02888 | -0.01712 | Gestational Age |
| Vaginal Delivery | PTB | -0.007 | 0.001 | -0.00896 | -0.00504 | Gestational Age |
| Mother, no high school degree | PTB | -0.003 | 0.004 | -0.01084 | 0.00484 | Gestational Age |
| Mother, high school degree only | PTB | -0.008 | 0.003 | -0.01388 | -0.00212 | Gestational Age |
| Mother, college degree | PTB | -0.008 | 0.003 | -0.01388 | -0.00212 | Gestational Age |
| Non-Hispanic White Mother | PTB | -0.011 | 0.001 | -0.01296 | -0.00904 | Gestational Age |
| Non-Hispanic Black Mother | PTB | -0.01 | 0.01 | -0.0296 | 0.0096 | Gestational Age |
| Unknown Father Age | PTB | -0.03 | 0.003 | -0.03588 | -0.02412 | Gestational Age |
| Known Father Age | PTB | -0.007 | 0.001 | -0.00896 | -0.00504 | Gestational Age |
| First Birth | PTB | -0.003 | 0.002 | -0.00692 | 0.00092 | Gestational Age |
| Prior Birth, none preterm | PTB | -0.009 | 0.002 | -0.01292 | -0.00508 | Gestational Age |
| Prior Preterm Birth | PTB | -0.023 | 0.005 | -0.0328 | -0.0132 | Gestational Age |
| Mother is Not Married | PTB | -0.016 | 0.002 | -0.01992 | -0.01208 | Gestational Age |
| Mother is Married | PTB | -0.008 | 0.001 | -0.00996 | -0.00604 | Gestational Age |
| Gained <20 lb | PTB | 0.001 | 0.005 | -0.0088 | 0.0108 | Gestational Age |
| Gained 21-30 lb | PTB | -0.008 | 0.003 | -0.01388 | -0.00212 | Gestational Age |
| Gained 31-40 lb | PTB | -0.009 | 0.005 | -0.0188 | 0.0008 | Gestational Age |
| Gained 40+ lb | PTB | -0.025 | 0.007 | -0.03872 | -0.01128 | Gestational Age |
| Teen (<= 19) | PTB | -0.009 | 0.005 | -0.0188 | 0.0008 | Gestational Age |
| High-Risk Age (>= 35) | PTB | -0.026 | 0.006 | -0.03776 | -0.01424 | Gestational Age |
| Age 20-34 | PTB | -0.009 | 0.001 | -0.01096 | -0.00704 | Gestational Age |
| No Birth Attendant | NTB | 0.015 | 0.004 | 0.00716 | 0.02284 | Gestational Age |
| Birth Attended by Provider | NTB | 0.018 | 0.003 | 0.01212 | 0.02388 | Gestational Age |
| Did Not Smoke During Pregnancy | NTB | 0.013 | 0.004 | 0.00516 | 0.02084 | Gestational Age |

| | | | | | | |
|---|---|---|---|---|---|---|
| Smoked During Pregnancy | NTB | 0.031 | 0.005 | 0.0212 | 0.0408 | Gestational Age |
| Female Child | NTB | 0.012 | 0.003 | 0.00612 | 0.01788 | Gestational Age |
| Male Child | NTB | 0.02 | 0.004 | 0.01216 | 0.02784 | Gestational Age |
| Cesarean Delivery | NTB | 0.031 | 0.004 | 0.02316 | 0.03884 | Gestational Age |
| Vaginal Delivery | NTB | 0.011 | 0.004 | 0.00316 | 0.01884 | Gestational Age |
| Mother, no high school degree | NTB | 0.012 | 0.004 | 0.00416 | 0.01984 | Gestational Age |
| Mother, high school degree only | NTB | 0.005 | 0.005 | -0.0048 | 0.0148 | Gestational Age |
| Mother, college degree | NTB | 0.017 | 0.006 | 0.00524 | 0.02876 | Gestational Age |
| Non-Hispanic White Mother | NTB | 0.016 | 0.004 | 0.00816 | 0.02384 | Gestational Age |
| Non-Hispanic Black Mother | NTB | 0.011 | 0.013 | -0.01448 | 0.03648 | Gestational Age |
| Unknown Father Age | NTB | 0.036 | 0.007 | 0.02228 | 0.04972 | Gestational Age |
| Known Father Age | NTB | 0.012 | 0.003 | 0.00612 | 0.01788 | Gestational Age |
| First Birth | NTB | 0.012 | 0.005 | 0.0022 | 0.0218 | Gestational Age |
| Prior Birth, none preterm | NTB | 0.009 | 0.003 | 0.00312 | 0.01488 | Gestational Age |
| Prior Preterm Birth | NTB | 0.031 | 0.01 | 0.0114 | 0.0506 | Gestational Age |
| Mother is Not Married | NTB | 0.023 | 0.003 | 0.01712 | 0.02888 | Gestational Age |
| Mother is Married | NTB | 0.012 | 0.004 | 0.00416 | 0.01984 | Gestational Age |
| Gained <20 lb | NTB | -0.006 | 0.006 | -0.01776 | 0.00576 | Gestational Age |
| Gained 21-30 lb | NTB | 0.014 | 0.006 | 0.00224 | 0.02576 | Gestational Age |
| Gained 31-40 lb | NTB | 0.022 | 0.003 | 0.01612 | 0.02788 | Gestational Age |
| Gained 40+ lb | NTB | 0.031 | 0.009 | 0.01336 | 0.04864 | Gestational Age |
| Teen (<= 19) | NTB | 0.017 | 0.009 | -0.00064 | 0.03464 | Gestational Age |
| High-Risk Age (>= 35) | NTB | 0.023 | 0.015 | -0.0064 | 0.0524 | Gestational Age |
| Age 20-34 | NTB | 0.014 | 0.001 | 0.01204 | 0.01596 | Gestational Age |
| No Birth Attendant | NBW | 0 | 0.005 | -0.0098 | 0.0098 | Birthweight |
| Birth Attended by Provider | NBW | 0.006 | 0.002 | 0.00208 | 0.00992 | Birthweight |
| Did Not Smoke During Pregnancy | NBW | 0.003 | 0.001 | 0.00104 | 0.00496 | Birthweight |
| Smoked During Pregnancy | NBW | 0.009 | 0.002 | 0.00508 | 0.01292 | Birthweight |
| Female Child | NBW | 0.006 | 0.003 | 0.00012 | 0.01188 | Birthweight |
| Male Child | NBW | 0.001 | 0.003 | -0.00488 | 0.00688 | Birthweight |
| Cesarean Delivery | NBW | -0.002 | 0.007 | -0.01572 | 0.01172 | Birthweight |
| Vaginal Delivery | NBW | 0.006 | 0.003 | 0.00012 | 0.01188 | Birthweight |
| Mother, no high school degree | NBW | -0.004 | 0.005 | -0.0138 | 0.0058 | Birthweight |
| Mother, high school degree only | NBW | 0.001 | 0.005 | -0.0088 | 0.0108 | Birthweight |
| Mother, college degree | NBW | 0.013 | 0.003 | 0.00712 | 0.01888 | Birthweight |
| Non-Hispanic White Mother | NBW | 0.004 | 0.002 | 8E-05 | 0.00792 | Birthweight |
| Non-Hispanic Black Mother | NBW | 0.006 | 0.008 | -0.00968 | 0.02168 | Birthweight |
| Unknown Father Age | NBW | 0.006 | 0.002 | 0.00208 | 0.00992 | Birthweight |
| Known Father Age | NBW | 0.004 | 0.001 | 0.00204 | 0.00596 | Birthweight |
| First Birth | NBW | -0.006 | 0.003 | -0.01188 | -0.00012 | Birthweight |
| Prior Birth, none preterm | NBW | 0.012 | 0.004 | 0.00416 | 0.01984 | Birthweight |
| Prior Preterm Birth | NBW | 0.004 | 0.007 | -0.00972 | 0.01772 | Birthweight |
| Mother is Not Married | NBW | 0.004 | 0.005 | -0.0058 | 0.0138 | Birthweight |

| | | | | | | |
|---|---|---|---|---|---|---|
| Mother is Married | NBW | 0.004 | 0.002 | 8E-05 | 0.00792 | Birthweight |
| Gained <20 lb | NBW | -0.012 | 0.006 | -0.02376 | -0.00024 | Birthweight |
| Gained 21-30 lb | NBW | 0.003 | 0.006 | -0.00876 | 0.01476 | Birthweight |
| Gained 31-40 lb | NBW | 0.005 | 0.004 | -0.00284 | 0.01284 | Birthweight |
| Gained 40+ lb | NBW | 0.023 | 0.005 | 0.0132 | 0.0328 | Birthweight |
| Teen (<= 19) | NBW | 0.002 | 0.008 | -0.01368 | 0.01768 | Birthweight |
| High-Risk Age (>= 35) | NBW | 0.014 | 0.011 | -0.00756 | 0.03556 | Birthweight |
| Age 20-34 | NBW | 0.002 | 0.002 | -0.00192 | 0.00592 | Birthweight |
| No Birth Attendant | LTB | -0.004 | 0.005 | -0.0138 | 0.0058 | Gestational Age |
| Birth Attended by Provider | LTB | -0.004 | 0.001 | -0.00596 | -0.00204 | Gestational Age |
| Did Not Smoke During Pregnancy | LTB | -0.004 | 0.002 | -0.00792 | -8E-05 | Gestational Age |
| Smoked During Pregnancy | LTB | -0.005 | 0.005 | -0.0148 | 0.0048 | Gestational Age |
| Female Child | LTB | -0.003 | 0.003 | -0.00888 | 0.00288 | Gestational Age |
| Male Child | LTB | -0.006 | 0.001 | -0.00796 | -0.00404 | Gestational Age |
| Cesarean Delivery | LTB | -0.003 | 0.003 | -0.00888 | 0.00288 | Gestational Age |
| Vaginal Delivery | LTB | -0.005 | 0.002 | -0.00892 | -0.00108 | Gestational Age |
| Mother, no high school degree | LTB | -0.01 | 0.003 | -0.01588 | -0.00412 | Gestational Age |
| Mother, high school degree only | LTB | 0 | 0.002 | -0.00392 | 0.00392 | Gestational Age |
| Mother, college degree | LTB | -0.003 | 0.001 | -0.00496 | -0.00104 | Gestational Age |
| Non-Hispanic White Mother | LTB | -0.005 | 0.001 | -0.00696 | -0.00304 | Gestational Age |
| Non-Hispanic Black Mother | LTB | -0.002 | 0.005 | -0.0118 | 0.0078 | Gestational Age |
| Unknown Father Age | LTB | -0.006 | 0.003 | -0.01188 | -0.00012 | Gestational Age |
| Known Father Age | LTB | -0.004 | 0.001 | -0.00596 | -0.00204 | Gestational Age |
| First Birth | LTB | -0.014 | 0.003 | -0.01988 | -0.00812 | Gestational Age |
| Prior Birth, none preterm | LTB | 0.003 | 0.002 | -0.00092 | 0.00692 | Gestational Age |
| Prior Preterm Birth | LTB | -0.003 | 0.004 | -0.01084 | 0.00484 | Gestational Age |
| Mother is Not Married | LTB | -0.007 | 0.001 | -0.00896 | -0.00504 | Gestational Age |
| Mother is Married | LTB | -0.003 | 0.002 | -0.00692 | 0.00092 | Gestational Age |
| Gained <20 lb | LTB | 0.001 | 0.003 | -0.00488 | 0.00688 | Gestational Age |
| Gained 21-30 lb | LTB | -0.003 | 0.003 | -0.00888 | 0.00288 | Gestational Age |
| Gained 31-40 lb | LTB | -0.008 | 0.003 | -0.01388 | -0.00212 | Gestational Age |
| Gained 40+ lb | LTB | -0.007 | 0.003 | -0.01288 | -0.00112 | Gestational Age |
| Teen (<= 19) | LTB | -0.009 | 0.003 | -0.01488 | -0.00312 | Gestational Age |
| High-Risk Age (>= 35) | LTB | 0.004 | 0.004 | -0.00384 | 0.01184 | Gestational Age |
| Age 20-34 | LTB | -0.005 | 0.001 | -0.00696 | -0.00304 | Gestational Age |
| No Birth Attendant | LBW | 0.005 | 0.005 | -0.0048 | 0.0148 | Birthweight |
| Birth Attended by Provider | LBW | 0.003 | 0.001 | 0.00104 | 0.00496 | Birthweight |
| Did Not Smoke During Pregnancy | LBW | 0.004 | 0.002 | 8E-05 | 0.00792 | Birthweight |
| Smoked During Pregnancy | LBW | -0.002 | 0.005 | -0.0118 | 0.0078 | Birthweight |
| Female Child | LBW | 0.003 | 0.002 | -0.00092 | 0.00692 | Birthweight |
| Male Child | LBW | 0.004 | 0.002 | 8E-05 | 0.00792 | Birthweight |
| Cesarean Delivery | LBW | 0.001 | 0.004 | -0.00684 | 0.00884 | Birthweight |
| Vaginal Delivery | LBW | 0.004 | 0.002 | 8E-05 | 0.00792 | Birthweight |

| | | | | | | |
|---|---|---|---|---|---|---|
| Mother, no high school degree | LBW | 0.005 | 0.002 | 0.00108 | 0.00892 | Birthweight |
| Mother, high school degree only | LBW | 0.003 | 0.002 | -0.00092 | 0.00692 | Birthweight |
| Mother, college degree | LBW | 0.004 | 0.002 | 8E-05 | 0.00792 | Birthweight |
| Non-Hispanic White Mother | LBW | 0.004 | 0.001 | 0.00204 | 0.00596 | Birthweight |
| Non-Hispanic Black Mother | LBW | 0 | 0.007 | -0.01372 | 0.01372 | Birthweight |
| Unknown Father Age | LBW | 0.004 | 0.004 | -0.00384 | 0.01184 | Birthweight |
| Known Father Age | LBW | 0.003 | 0.001 | 0.00104 | 0.00496 | Birthweight |
| First Birth | LBW | 0.002 | 0.002 | -0.00192 | 0.00592 | Birthweight |
| Prior Birth, none preterm | LBW | 0.004 | 0.002 | 8E-05 | 0.00792 | Birthweight |
| Prior Preterm Birth | LBW | 0.004 | 0.003 | -0.00188 | 0.00988 | Birthweight |
| Mother is Not Married | LBW | 0.002 | 0.003 | -0.00388 | 0.00788 | Birthweight |
| Mother is Married | LBW | 0.003 | 0.001 | 0.00104 | 0.00496 | Birthweight |
| Gained <20 lb | LBW | 0.015 | 0.003 | 0.00912 | 0.02088 | Birthweight |
| Gained 21-30 lb | LBW | 0 | 0.004 | -0.00784 | 0.00784 | Birthweight |
| Gained 31-40 lb | LBW | -0.001 | 0.004 | -0.00884 | 0.00684 | Birthweight |
| Gained 40+ lb | LBW | 0 | 0.005 | -0.0098 | 0.0098 | Birthweight |
| Teen (<= 19) | LBW | 0.003 | 0.004 | -0.00484 | 0.01084 | Birthweight |
| High-Risk Age (>= 35) | LBW | -0.005 | 0.005 | -0.0148 | 0.0048 | Birthweight |
| Age 20-34 | LBW | 0.004 | 0.001 | 0.00204 | 0.00596 | Birthweight |
| No Birth Attendant | HBW | -0.004 | 0.007 | -0.01772 | 0.00972 | Birthweight |
| Birth Attended by Provider | HBW | -0.007 | 0.002 | -0.01092 | -0.00308 | Birthweight |
| Did Not Smoke During Pregnancy | HBW | -0.006 | 0.002 | -0.00992 | -0.00208 | Birthweight |
| Smoked During Pregnancy | HBW | -0.001 | 0.006 | -0.01276 | 0.01076 | Birthweight |
| Female Child | HBW | -0.005 | 0.002 | -0.00892 | -0.00108 | Birthweight |
| Male Child | HBW | -0.005 | 0.003 | -0.01088 | 0.00088 | Birthweight |
| Cesarean Delivery | HBW | 0.002 | 0.004 | -0.00584 | 0.00984 | Birthweight |
| Vaginal Delivery | HBW | -0.008 | 0.002 | -0.01192 | -0.00408 | Birthweight |
| Mother, no high school degree | HBW | 0.002 | 0.006 | -0.00976 | 0.01376 | Birthweight |
| Mother, high school degree only | HBW | -0.005 | 0.005 | -0.0148 | 0.0048 | Birthweight |
| Mother, college degree | HBW | -0.014 | 0.004 | -0.02184 | -0.00616 | Birthweight |
| Non-Hispanic White Mother | HBW | -0.006 | 0.002 | -0.00992 | -0.00208 | Birthweight |
| Non-Hispanic Black Mother | HBW | -0.002 | 0.004 | -0.00984 | 0.00584 | Birthweight |
| Unknown Father Age | HBW | -0.005 | 0.002 | -0.00892 | -0.00108 | Birthweight |
| Known Father Age | HBW | -0.006 | 0.002 | -0.00992 | -0.00208 | Birthweight |
| First Birth | HBW | 0.002 | 0.002 | -0.00192 | 0.00592 | Birthweight |
| Prior Birth, none preterm | HBW | -0.013 | 0.003 | -0.01888 | -0.00712 | Birthweight |
| Prior Preterm Birth | HBW | -0.004 | 0.004 | -0.01184 | 0.00384 | Birthweight |
| Mother is Not Married | HBW | -0.001 | 0.004 | -0.00884 | 0.00684 | Birthweight |
| Mother is Married | HBW | -0.007 | 0.002 | -0.01092 | -0.00308 | Birthweight |
| Gained <20 lb | HBW | -0.005 | 0.004 | -0.01284 | 0.00284 | Birthweight |
| Gained 21-30 lb | HBW | 0.001 | 0.003 | -0.00488 | 0.00688 | Birthweight |
| Gained 31-40 lb | HBW | 0 | 0.003 | -0.00588 | 0.00588 | Birthweight |
| Gained 40+ lb | HBW | -0.025 | 0.004 | -0.03284 | -0.01716 | Birthweight |

| | | | | | | |
|---|---|---|---|---|---|---|
| Teen (<= 19) | HBW | 0.002 | 0.006 | -0.00976 | 0.01376 | Birthweight |
| High-Risk Age (>= 35) | HBW | -0.014 | 0.009 | -0.03164 | 0.00364 | Birthweight |
| Age 20-34 | HBW | -0.005 | 0.003 | -0.01088 | 0.00088 | Birthweight |
